# Sub-second and multi-second dopamine dynamics underlie variability in human time perception

**DOI:** 10.1101/2024.02.09.24302276

**Authors:** Renata Sadibolova, Emily K. DiMarco, Angela Jiang, Benjamin Maas, Stephen B. Tatter, Adrian Laxton, Kenneth T. Kishida, Devin B. Terhune

**Author notes:** Co-senior authors (Kenneth T. Kishida, Devin B. Terhune). **One-Sentence Summary:** Tonic and phasic dopamine fluctuations in striatum differentially relate to variations in human time perception.

## Abstract

Timing behaviour and the perception of time are fundamental to cognitive and emotional processes in humans. In non-human model organisms, the neuromodulator dopamine has been associated with variations in timing behaviour, but the connection between variations in dopamine levels and the human experience of time has not been directly assessed. Here, we report how dopamine levels in human striatum, measured with sub-second temporal resolution during awake deep brain stimulation surgery, relate to participants’ perceptual judgements of time intervals. Fast, phasic, dopaminergic signals were associated with underestimation of temporal intervals, whereas slower, tonic, decreases in dopamine were associated with poorer temporal precision. Our findings suggest a delicate and complex role for the dynamics and tone of dopaminergic signals in the conscious experience of time in humans.

---

The decisions we make, the memories we retain, and the way we perceive our surroundings are all intrinsically tied to our experience of time (*1–4*). Therefore, investigations into human cognition and consciousness require consideration of the mechanisms governing time perception, particularly in a time frame on the order of milliseconds to seconds which is integral to basic perceptual and cognitive processes (*5–8*).

The neurobiological mechanisms underlying the timing of these relatively short intervals remain elusive. Past evidence implicates the striatum and striatal dopamine in time perception (*9–17*), with higher dopamine levels associated with a tendency to perceive short intervals of time as longer (*18–22*). This association was drawn largely on the basis of pharmacological experiments and work in non-human animal models (*9–13*), owing to the inherent challenges of collecting direct dopamine measurements in the living human brain during conscious behaviour (*23*, *24*). However, recent optogenetic work in rodents has introduced a new perspective by demonstrating that rapid changes in dopamine signals (i.e., bursts in phasic dopaminergic neuron activity) may induce temporal underestimation (*25*). This finding challenges classic timing models that are based on relatively slow response pharmacological effects (*18–22*, *26*). Notably, the contrast between these lines of evidence isn’t just in their resulting interpretation, but also for the very different timescales by which the dopaminergic system was observed to be exerting its influence.

Here we report the first human fast scan cyclic voltammetry (FSCV) (*23*, *24*) study to directly assess the role of sub-second striatal dopamine signals in human time perception. FSCV during the implantation of deep brain stimulation electrodes for the treatment of Parkinson’s disease (PD) symptoms (*27*) enables the recording of both tonic and phasic striatal dopamine with 100ms precision during concurrent behavioural assessments (*23*, *24*, *28–33*). Tonic dopamine refers to the sustained signaling over the course of minutes whereas fast, phasic, dopamine fluctuations occur rapidly within tens to hundreds of milliseconds (*34*). The present study tests the hypothesis that interval timing is differentially related to dopaminergic activity at varying timescales (*34–36*), as observed in non-timing cognitive functions (*37*). Additionally, our approach allows for simultaneous and colocalized measurements of serotonin (*28*, *30*) to assess the neurochemical specificity of any observed effects.

Seventeen healthy controls from a separate study (*38*) and six patients undergoing deep brain stimulation (DBS) surgery completed a visual temporal bisection task (*39*). All participants first learned two anchor intervals (500ms and 1,100ms) before judging whether specific intervals (of varying durations between 500ms and 1,100ms) were closer in duration to the short or long anchor interval. This enabled an assessment of temporal accuracy and precision, defined by the degree of systematic error in time judgements and the sensitivity to interval differences, respectively. Concurrently, utilizing a safe and validated human-adapted FSCV protocol (*24*, *28*, *30–33*), we recorded in-vivo electrochemical responses at 10 Hz temporal resolution from patients’ caudate (Fig. S1). To estimate dopamine and serotonin concentrations, we trained elastic net penalised regression models using in-vitro calibration data following prior work (*24*, *28*, *30*–*33,* also see Supplemental Information). Subsequently, we quantified both short-lived transient (i.e., phasic) changes and slowly changing (i.e., tonic, at ∼4 min timescale) concentrations to investigate the association between these neurochemical signals and changes in participants’ temporal accuracy and precision across trials.

## Elevated *phasic* dopamine concentrations are associated with temporal underestimation

We tested the hypothesis that elevated striatal dopamine transients are associated with increased temporal underestimation (*25*). A cluster analysis (*40*) revealed that short-interval temporal judgements were associated with phasic increases in dopamine levels at 625ms to 670ms after stimulus onset (Fig. 1C), *p*<.050. Notably, this association was not observed when comparing across objectively short and long stimulus durations and no effects were found for serotonin (Fig 1A,B,D, Supplementary Materials). Additional analyses show that the likelihood of short judgements is significantly increased during higher mean single-trial dopamine concentrations, but not serotonin concentrations, in the 625ms to 670ms time window after the stimulus onset (dopamine: *β*=-.11, *p*=.048, serotonin: *β*=-.06, *p*=.28).

**Fig. 1:**
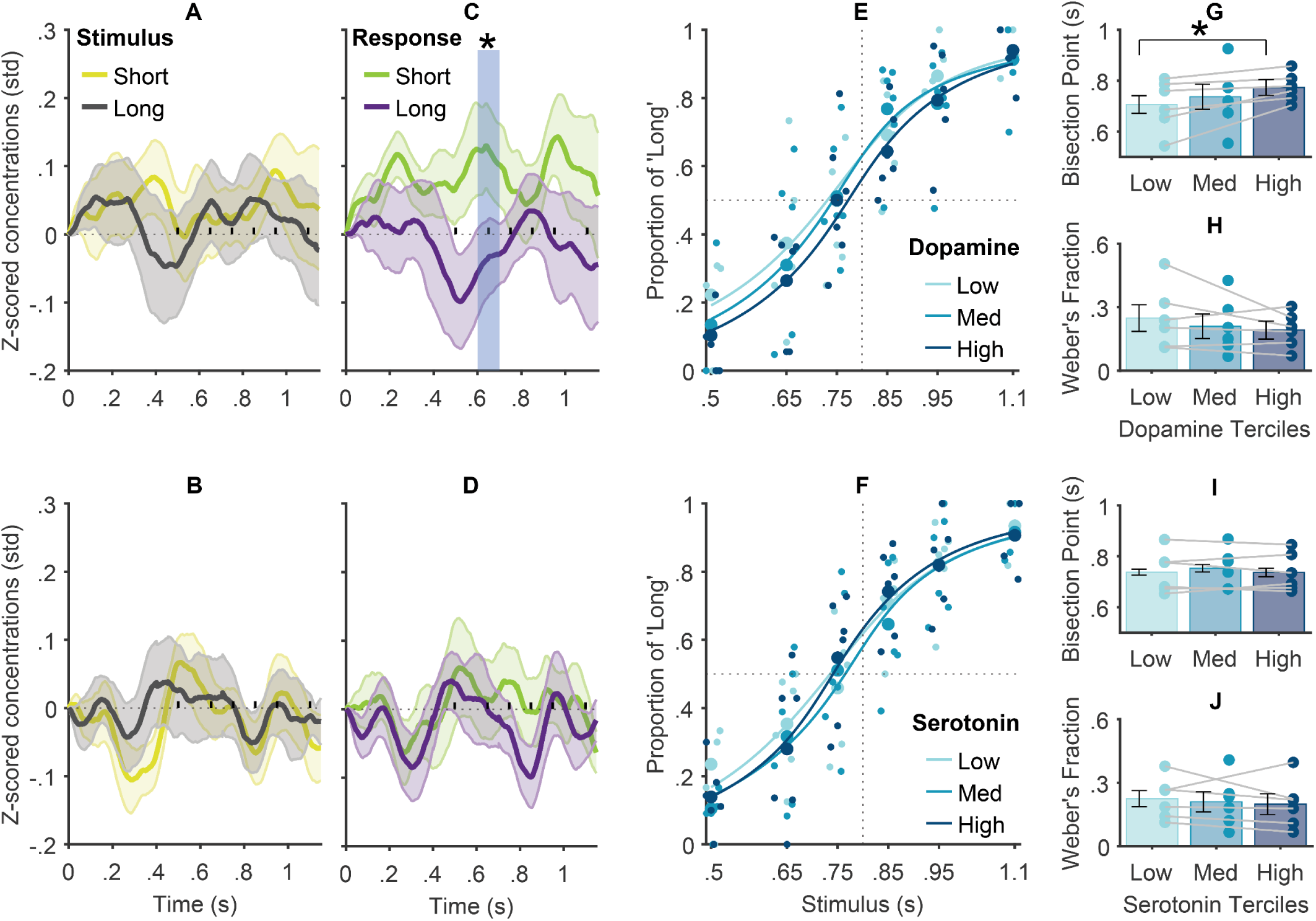
Relationships between phasic caudal changes in dopamine and serotonin levels and stimulus and response parameters. (A-D): Normalized dopamine (A,C) and serotonin (B,D) concentrations from stimulus onset as a function of (A-B) stimulus intervals and (C-D) responses. Shaded areas reflect standard error (SE); x-axis markers on the zero y-axis represent stimulus interval offsets, and the significant cluster*. (E-F) Psychometric functions (PF) fitted to the proportion of ‘long’ responses across stimulus intervals as a function of phasic dopamine (E) and serotonin (F) terciles (Low, Medium, and High). Markers represent individual patient datapoints; dotted lines mark accurate performance with 50% long and short responses (Point of subjective equality [PSE]=0.5) for the true stimulus mid-interval = 0.8 s. (G-J) Indices of the PFs fitted to individual patients’ data. Subjective mid-intervals (Bisection Points) and temporal precision (Weber’s fractions) across dopamine (G-H) and serotonin (I-J) terciles. Markers represent individual patient datapoints. Lines show for each patient the difference in time perception performance as a function of Dopamine or Serotonin tercile. Lower Bisection Point and Weber fraction values denote greater underestimation bias and temporal precision, respectively. * *p*<.050

To delve further into the relationship between phasic dopamine fluctuations and variations in temporal accuracy and precision, we partitioned trials (*n_(total)_*=300) for each patient according to the average dopamine level in the cluster window into low, medium, and high terciles. Using generalized mixed-effects modelling (*41*), we compared the temporal psychometric functions across these terciles (Fig. 1E-F), where left-and right-ward shifts signify under-and over-estimation biases, respectively, and steeper functions signify superior precision. Consistent with both the foregoing analyses and rodent data (*25*), our results point to a significant underestimation bias concurrent with higher dopamine transients (Fig. 1E,G; *β*=-.15, *p*=.010). This effect was not observed for serotonin (Fig. 1F,I; *β*=-.01, *p*=.82). Finally, we complemented our analyses with Bayesian assessment of effect prevalence (Fig. S2) which is uniquely suited for experiments with small sample sizes and large trial numbers such as ours (*42*). The results suggest an 83% probability (with at least 43% at the lower boundary) of detecting statistically significant classification of temporal judgments from dopamine signals within 1,100ms from stimulus onset if our methods are replicated (the lower boundary for prevalence estimate reduces to 33% under more stringent assessment).

Although the observed effects reached statistical significance at 625ms to 670ms from the stimulus onset, our data show that dopamine timeseries for short and long judgments began diverging earlier at around 500ms (short anchor duration) until approximately 800ms (mid-interval of presented stimulus range) (Fig. 1C). This suggests that dopamine neurons may anticipate the impending stimulus offset during this period, which would facilitate efficient decision-making by favouring the selection of the short reference anchor and decreasing the likelihood of classifying the response as ’long’ when the offset falls within this timeframe. Critically, this plausibly explains why dopamine responses in this time window differed between short and long subjective responses but not following the objective short and long stimulus intervals (Fig. 1A). In line with this interpretation, lower dopamine responses for long judgements might partly reflect temporal discounting of the reward value (*43*). Notably, in some population clock models, dopaminergic projections modify cortical population dynamics through processes linked to reward prediction error (*44*), implying that a transient increase in dopamine may lead to temporal underestimation by decelerating trajectories of population dynamics (*45*, *46*). Though, evidence from research in rodents suggests that dopamine neurons exert control over temporal judgments independently of reward processing (*25*), highlighting the timing specificity of the striatal dopaminergic system as an alternative interpretation. More research is required to further distinguish between these competing hypotheses.

### Elevated *tonic* dopamine concentrations underlie superior temporal precision

Parkinson’s Disease is marked by poorer temporal precision (*47–50*). Owing to the depletion of dopamine neurons in this condition (*51*), it has been hypothesized that striatal dopamine concentrations scale with temporal precision. This has been corroborated in pharmacological research (*17*) but, to our knowledge, has not yet been shown directly with in vivo dopamine measurement in humans. Consistent with this hypothesis, we observed diminished temporal precision in patients (Just Noticeable Difference, JND=.14) compared to controls (JND=.09) (Fig. 2A), *p*=.017, although the Bayesian evidence for this effect was ambiguous, *BF_10_*=1.03. Further, we found that patients’ slowly changing tonic dopamine levels over the course of completing the task, predicted variation in their temporal precision (*β*=.50, *p*=.009, *BF_10_*=1.70), such that elevated tonic dopamine levels were associated with improved precision (Fig. 2D,F). This effect appeared to be specific to tonic dopamine, as we found evidence supporting the corresponding null hypothesis for serotonin (Fig. 2G,I, *β*=.26, *p*=.35, *BF_10_*=.09). Furthermore, exploratory analyses on phasic dopamine and serotonin transients did not suggest a link with temporal precision (Fig. 1E-J; dopamine: *β*=.49, *p*=.39, serotonin: *β*=.61, *p*=.13).

**Fig. 2:**
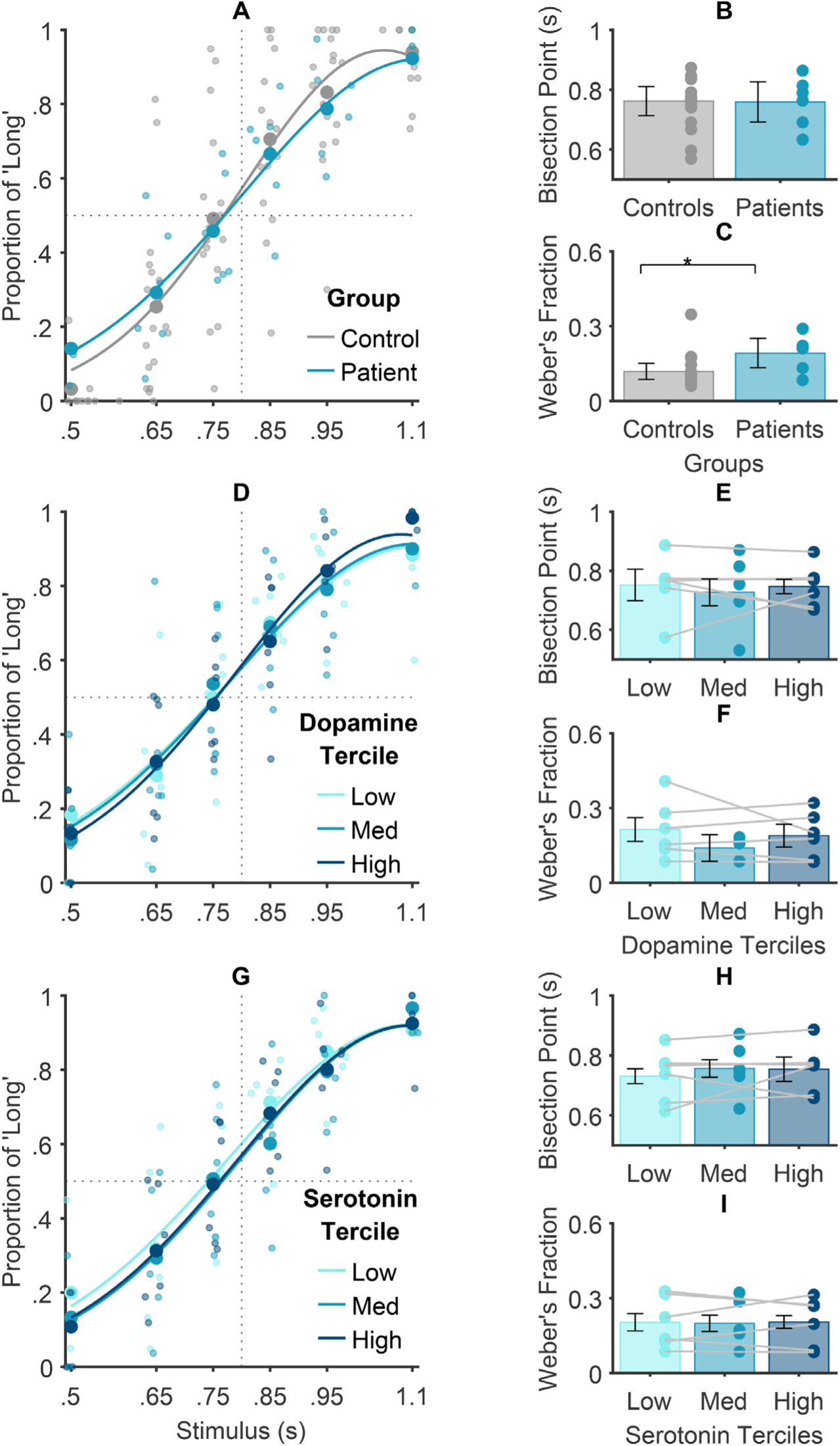
Relationships between tonic caudal changes in dopamine and serotonin levels and time perception performance. (A, D, G) PFs were fitted to the proportion of ‘Long’ responses across stimulus intervals. The horizontal dotted line indicates the point of indifference, i.e., 50% of ‘long’ and 50% of ‘short’ responses. The vertical dotted lines denote the absence of a perceptual bias, such that relative leftward and rightward PF shifts represent the over-and under-estimation biases, respectively. PFs show the fit to (A) patient and control group data, (B) across patients’ dopamine terciles, and (C) patients’ serotonin terciles. (B-C,E-F,H-I) Indices of the PFs fitted to individual patients’ and controls’ data. Subjective mid-intervals (Bisection Points) and temporal precision (Weber’s fractions) across groups (B-C), and patients’ dopamine (E-F) and serotonin (H-I) terciles. Lines show for each patient the difference in time perception performance as a function of Dopamine or Serotonin tercile. Lower Bisection Points (leftward PF shift) and Weber’s fractions (steeper PF slopes) denote increasing underestimation bias and temporal precision, respectively. Markers reflect individual participant datapoints.

Future research investigating the interplay between across-trial interval learning and tonic dopamine could shed further light on the mechanisms underlying our observed association between tonic dopamine and temporal precision. For example, decreased temporal precision may relate to observed lower motivational states being associated with reduced dopamine levels (*52*). Our observations may also reflect the role of tonic dopamine in behaviour reinforcement, where a hypoactive tonic firing rate, demonstrated to hinder the extinction of previously reinforced behaviours (*53*) could contribute to the observed bias towards responses from prior trials in our study. This bias could lead to increased response variability across trials, subsequently diminishing temporal precision.

Our final set of analyses sought to discriminate between two contrasting hypotheses regarding the role of *tonic* striatal dopamine in temporal accuracy. The first hypothesis, grounded in pharmacological research (*18–22*), predicts a positive association between dopamine and temporal accuracy whereas the second hypothesis predicts no association because of the absence of clear evidence for atypical temporal accuracy in dopamine-depleted PD (*54*). In alignment with the latter, our observations revealed comparable accuracy across patients and controls (Fig. 2A,B), *p*=.76, with Bayesian evidence for the null hypothesis, *BF_10_*=.09. This observation was strengthened by the lack of correspondence between temporal accuracy and variations observed in patients’ tonic dopamine levels (comprising a ∼4 min period) over the course of experimental sessions (Fig. 2D,E; *p*=.89, *BF_10_*=.06). This was consistent across different analysis window lengths, underscoring that temporal accuracy is not related to steady-state (i.e., slowly changing or tonic) striatal dopamine levels. Similarly, no link was found between temporal accuracy and tonic serotonin fluctuations (Fig. 2G,H; *p*=.24, *BF_10_*=.11). These observations challenge internal clock models proposing that higher dopamine levels produce a tendency to perceive time as lasting longer (*20*). The pharmacological evidence used to support these models (*18–22*) plausibly reflects mechanisms other than a putative internal clock with its speed of ticking purportedly controlled by striatal dopamine levels. Perhaps most compellingly, our observations reveal differential effects of phasic and tonic dopamine dynamics on temporal accuracy. This suggests a far more intricate relationship between the striatal dopamine system and time perception that warrants further exploration and refinement of existing theoretical frameworks.

Collectively, our observations indicate that changes in tonic dopamine levels may specifically underlie the precision of temporal judgments, whereas we did not observe compelling evidence linking temporal precision to phasic dopamine nor to phasic or tonic serotonin. Coupled with our finding showing that phasic dopamine increases were selectively associated with a higher underestimation bias, these results cumulatively suggest differential roles for tonic and phasic dopamine in human time perception.

### New insights into the role of dopamine in time perception

The present results offer a nuanced perspective on how striatal dopamine fluctuations underlie different features of human time perception. Grace (*36*) has proposed that dopamine dynamics operate across multiple timescales, encompassing a fast phasic release in response to stimuli and a slow tonic release, which sustains steady-state concentration levels and modulates phasic firing. In line with this perspective, other studies have shown that striatal tonic and phasic firing can yield diverse and even opposing effects on behaviour (*37*, *55*). Investigations into the role of striatal dopamine in time perception faces similar challenges due to conflicting observations linking elevated dopamine with both temporal overestimation (*18–22*) and underestimation (*25*, *56*). Here we leveraged advances in neurochemical measurement methods in human participants to show that heightened phasic bursts of striatal dopamine are uniquely associated with temporal underestimation whereas higher tonic dopamine concentrations correlated with superior temporal precision. Moreover, our results highlight the neurochemical specificity of these effects as our analyses suggest that the observed effects are specific to dopamine fluctuations and are not observed with corresponding variations in serotonin levels, which have been linked to interval timing in pharmacological studies (*57–59*).

Dopamine assumes a central role in brain function regulation and is implicated in a broad spectrum of disorders, including Parkinson’s disease (*51*). These roles, rooted in dopamine’s evolutionary antiquity, persist across species (*60*). Recent shifts in dopamine transmission models have recognized the importance of spatiotemporal precision in certain dopamine functions. A specialized architecture has been proposed, involving release-receptor assemblies at micrometre scales, to account for the distinct functions associated with the tonic concentrations and transient bursts of activity of dopaminergic neurons (*34*). This theoretical framework emphasizes a more refined spatiotemporal resolution of dopamine signalling to mediate diversity of functions within the striatal dopamine system. Ongoing research investigating the principles of these theories may profoundly impact our understanding of past evidence linking a wide range of phenomena to the function of the striatal dopamine system, including time perception.

Studies employing FSCV in animal models have shed light on how dopaminergic transmission and dopamine dynamics contribute to the neurochemical mechanisms of drug addiction, PD, and schizophrenia (*61*). Nonetheless, until recently, such research had not been feasible in the human brain, owing to various methodological hurdles (*23*, *24*, *62*). Our results suggest new directions for research that harness recent methodological advances in the measurement of neurochemical dynamics (*33*, *63*) and their function in time perception and related behaviour in humans. Advancing our understanding of the neurobiological foundations of human time perception will refine existing theories (*20*, *44*, *64*, *65*), impacting healthy and dysfunctional timing and the knowledge concerning the timing in human cognition more broadly. This includes implications for metacognition, reinforcement learning, reward processing, and understanding neurological and psychiatric conditions like schizophrenia, addiction disorders and impulse control that involve aberrant dopamine functioning (*66*, *67*). For example, recent research indicates that heterogeneity in PD symptoms may be accounted for by differences in time perception, with implications for understanding dopaminergic mechanisms in time perception and PD symptomatology (*38*). These insights are crucial for developing nuanced treatment approaches and enhancing our comprehension of the complex interplay between dopamine, time perception, and human cognition. Consequently, human FSCV emerges as a valuable set of methods for investigating the neurochemical underpinnings of human time perception and germane cognitive functions.

## Data Availability

Data is available upon reasonable request.

## Author contributions

Conceptualization: RS, EDM, KTK, DBT

Methodology: RS, EDM, AJ, KTK, DBT

Investigation: EDM, AJ, SBT, AL, KTK

Visualization: RS, BM

Funding acquisition: KTK, DBT

Project administration: KTK, DBT

Supervision: KTK, DBT

Writing – original draft: RS

Writing – review & editing: RS, EDM, SBT, AL, KTK, DBT

## Data and materials availability

All data and code will be released with a 1-year embargo from the publication date.

## Supplementary Materials

### Materials and Methods

The study comprised two sessions (1 to 10 days apart for the patients and the same day with a ten-minute break between sessions for controls), each including training and experimental phases. All participants achieved 65% accuracy in 20 consecutive training trials, with a single patient and a single control requiring, respectively, 48 and 30 trials in the first session and another patient requiring 21 trials in the second session. Control participants and patients not in the surgical setting (session 1) were seated at a desk approximately 70 cm from the monitor and completed four blocks of fifty experimental trials. During surgery (session 2), PD patients sat in a semi-upright position and viewed the monitor at a distance of approximately 100 cm. Patients were on their dopamine replacement medication in session 1 whereas their medication was withheld in session 2 at least 12 hours prior to their surgery (*27*). Our experimental protocol afforded up to 30 minutes for the experimental task with concurrent FSCV recording (*24*, *32*), during which PD patients completed six blocks of 50 experimental trials.

#### Participants

Six patients with Parkinson’s disease (PD) (2 females, 4 males) between the ages of 62 and 73 years (*M*_Age_=67.7, *SD*=4.5) participated in this study during DBS surgery. For behavioural analyses only, we additionally included a sample of 17 healthy controls (7 females, 10 males, aged 50-66; *M*_Age_=57.5, *SD*=5.0) from a prior study (*38*), with no psychiatric or neurological conditions. We excluded 3 patients with a psychiatric diagnosis. All participants provided informed written consent in accordance with approval by the IRB committee at Wake Forest University Health Sciences (IRB00017138 and IRB00044216).

#### Experimental task

Participants completed a visual temporal bisection task with stimulus presentation implemented with Psychtoolbox-3 (*68*) in MATLAB R2018b (MathWorks, Natick, USA). Each trial consisted of a jittered interstimulus interval (ISI1; blank black screen) drawn from a truncated Poisson distribution (400-600ms), a white circle (∼2° of visual field) that varied in duration (500, 650, 750, 850, 950, or 1100ms), a second interstimulus interval (ISI2; blank black screen; 900ms), and a two-alternative forced choice judgment prompt (“S”=short; “L”=long). The response prompt on screen (i.e., [S L] or [L S]) was randomized on every trial to counterbalance handedness and visual presentation effects. Participants responded with the left or right shoulder keys of a Logitech game controller (Logitech International S.A.), corresponding to the respective response letter location on the monitor. In the 20-trial training phase, participants learnt two anchor intervals (500 and 1100ms) comprising equal proportions of each stimulus. Additional training trials automatically followed if accuracy was below 65% until this target was reached in the lattermost 20 trials or until the maximum training time of 7 minutes had passed. In the subsequent experimental phase, participants were presented with white circles of varying intervals (500-1100ms) and judged whether they were closer in duration to the trained short or long anchor intervals. All six experimental (50-trial) blocks began with two reminders of each anchor stimulus (4 trials); the remaining 46 trials included 10 repetitions of the 4 middle stimuli and 3 repetitions of each anchor stimulus.

#### Fast-Scan Cyclic Voltammetry (FSCV)

Fast scan cyclic voltammetry (FSCV) was performed using carbon fibre microelectrodes surgically placed in the caudate (Fig. S1; 6 channels, 5 patients) or thalamus (Fig. S1; 1 channel, 1 patient, excluded to focus analyses on caudate region). Briefly, the FSCV measurement protocol is the same as previously reported in humans: hold working electrode at -0.6V for 90ms, ramp to +1.4 V and back to -0.6V at 400 V/s, and repeat, for an overall electrochemical sampling rate of 10 Hz frequency reported (*23*, *24*, *63*, *28–33*, *38*, *62*). The resulting raw electrochemical current was measured at a sampling rate of 250kHz, thus providing a raw 10ms voltammogram (2500 samples at 250KHz). These data were subjected to analysis as described previously (*24*, *28*) and below.

**Figure S1.**
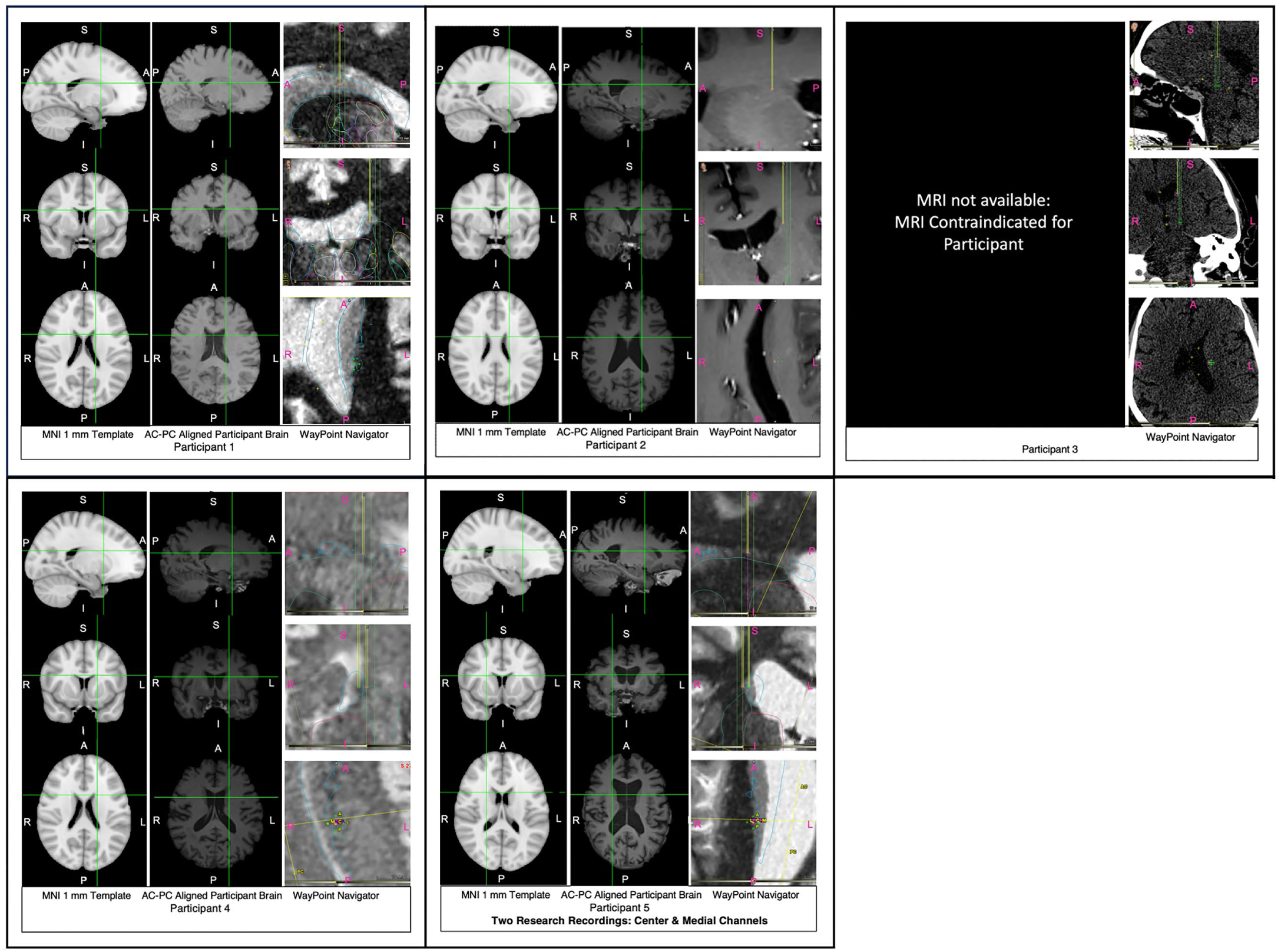
Electrode Coordinate Locations on MNI 152 Template, AC-PC aligned, and WayPoint Navigator Software. For each participant, from left to right, electrode coordinates are depicted on the MNI 152 1 mm template, AC-PC aligned participant brains, and WayPoint Navigator Surgical Planning Software Suite. Normalization to the MNI 152 template was achieved through FSL(*69*). Each figure panel shows sagittal, coronal, and axial planes for each participant from top to bottom. An exception is noted for Participant 3, for whom MRI was contraindicated and unavailable. For Participant 5, two distinct recordings were obtained from the same depth, one from a center channel, and another from a medial channel with a 2 mm offset. Abbreviations: AC-PC = anterior commissure-posterior commissure; S = superior; I = inferior; P = posterior; A = anterior; R = right; L = left; MRI = magnetic resonance imaging.

#### Analyses

#### Behavioural data analysis

Trials with premature responses (during ISI2) were excluded (*M*=1.50, *SD*=1.76, range: 0-5). We first tested whether psychophysical markers of time perception differed between controls and PD patients. Using the *MixedPsy* package (*41*) in R (*70*), we fitted a generalized linear mixed-effects model (GLMM) to the proportion of long responses, including the group and demeaned stimulus intervals and their interaction as fixed-effects parameters and stimulus interval slopes and intercepts for participants as random effects. Temporal accuracy (Point of Subjective Equality; PSE or Bisection Point; BP) and temporal precision (Just Noticeable Difference; JND, or Weber’s Fraction; WF) were derived, respectively, from the intercepts and slopes of a cumulative normal distribution (probit) function (*41*). Leftward (negative PSE) and rightward (positive PSE) shifts of this function reflect, respectively, temporal over-and under-estimation biases whereas a steeper function (lower JND) reflects greater temporal precision.

#### Neurochemical concentrations

We trained statistical models for optimal out-of-probe prediction of striatal DA and SE concentrations following prior work (*24*, *28*, *30*) and applied these models to estimate dopamine and serotonin concentrations from our human FSCV data. To this end, we produced the calibration data with known concentration labels following an in-vitro FSCV protocol described in detail elsewhere (*24*, *28*, *30*). Calibration data were collected using the same protocol as that used in the human experiments. Calibration data consisted of 10ms voltammograms recorded every 100ms (each voltammogram = 2500 samples at 250kHz), thus yielding 10Hz overall temporal resolution of DA and SE signal time series. We first sought to specify the set of parameters for optimal machine learning performance on naïve probes not included in the training. This was achieved by down-sampling calibration voltammograms by averaging the bins of every 30 samples for dopamine and 10 samples for serotonin. Each down-sampled voltammogram was concatenated with its first and second derivative with the resulting 249 (dopamine) and 747 (serotonin) samples submitted as predictors to an elastic-net regularized linear regression model (*71*, *72*). The outcome variable was a multivariate Gaussian distribution consisting of known concentration labels for pH, 3,4-dihydroxyphenylacetic acid (DOPAC), L-ascorbic acid (LAA) and dopamine (DA) or serotonin (SE). We trained separate models for DA and SE estimation using a 10-fold cross-validation procedure whilst searching for minimal mean squared error (unexplained variance) in a 2D penalty parameter space (penalty weight λ and a mixing term α, see Qian et al., 2013). The λ values were determined by an inbuilt function (*71*) and the α range was 0 to 1 in steps of 0.25. Our approach allowed for the reliable and generalized estimation of DA and SE concentrations against a background of varying pH activity as shown by model performance on two naïve calibration probes withheld from model-training (Root Mean Squared Error: dopamine = 0.632, serotonin = 0.113; Signal to Noise Ratio: dopamine = 3.99, serotonin = 18.91).

#### Phasic signal change and time perception

The final model estimates for patients’ in-vivo DA and SE concentrations were up-sampled by linear interpolation to 1000 Hz for the purposes of epoching. We extracted epochs relative to stimulus onset or stimulus offset and z-scored the signal (±3 sec relative to stimulus onset) to subtract away fluctuations at slower timescales. Insofar as the z-score normalization brings the epochs into a common frame of reference without affecting their individual shapes, we compared the transient within-trial signal changes both within and across participants.

Our next objective was to assess if patients’ binary response patterns in the temporal bisection task varied with transient changes in caudal DA and SE signals during and after stimulus presentation. We analysed timeseries aligned to stimulus onset [0 to 1400ms] and offset [-500 to 900ms]. We applied a cluster mass test procedure (*73*, *74*) for detecting the time window where neurochemical concentrations differed at the group level as a function of the stimulus interval, response (short or long), and their interaction, including the by-participant random error for the interaction term in the model. Clusters were identified with the threshold-free cluster enhancement (*tfce*) method (*75*) and assessed for significance with the *Rde_keradPajouh_renaud* (*76*) permutation method (10,000 iterations) in the *permuco* package (*40*) in R (*70*).

In the subsequent analyses, we submitted mean single-trial concentrations in identified cluster window(s) and stimulus interval (short vs. long) as fixed-effects predictors in a GLMM with binary responses (short vs. long) as the outcome variable and patient probes as the random intercept term. We additionally demarcated trials for individual probes into low, medium, and high DA and SE terciles of the mean signal distribution in the cluster window(s) and fitted GLMMs with signal tercile and stimulus predictors, proportion of long responses as the outcome variable, including the uncorrelated random intercepts and by-stimulus slopes for the probe. This allowed us to assess whether transient neurochemical fluctuations related to psychophysical indices of temporal accuracy and precision (*41*).

Although appropriate for its improved statistical power in dealing with multiple comparison tests (*40*, *73*), the cluster permutation analysis disregards random effects associated with participants and their interactions with fixed effects. Whereas the GLMM remedies some of those issues, we complemented these analyses with Bayesian assessments of the effect prevalence, which shifts the focus from population mean estimates to individuals and is particularly well-suited for experiments with small sample sizes and large trial numbers (*42*). For each PD patient, we trained 10-fold cross-validated logistic regression models in a sliding window across the signal time series (200ms width, steps of 20ms) with neurochemical concentrations as predictors and binary responses as the outcome. To ensure that the models were unbiased, the predictors were normalized, and the training data were balanced by subsampling trials from the response class with more trials, so that the number of trials was matched to the class with fewer trials. The minimal size of a training dataset in this study was N=212 trials. Area under the curve (AUC) was computed to assess classification performance with values of 0.50 and 1.00 representing chance and perfect classification, respectively. We applied one-tailed one-sample t-tests to compare AUC values (10 folds) against 50% chance performance at each timepoint for each participant. The alpha level was lowered to 0.01 to correct for multiple comparisons (46 time points); however, given that this is the first human study investigating in-vivo human sub-second neurochemical fluctuations relating to time perception, we additionally include the results without the multiple comparison correction (*28*). The Bayesian prevalence analysis provides the population prevalence estimate (maximum a posteriori) together with its associated uncertainty intervals (highest posterior density intervals [HPDI] or lower bound posterior quantiles). Expanding upon the traditional prevalence analysis, we also followed Ince et al.’s (*42*) approach to obtain prevalence estimates for different effect sizes.

#### Tonic signal change and time perception

Our previous analyses were complemented with further mixed-effects modelling to assess the role of tonic DA concentrations in time perception. To facilitate comparison across all patient probes, we z-scored the concentrations observed for ongoing stimuli using the mean and standard deviation of these concentrations across all trials. We subsequently computed baseline-corrected mean concentrations for individual trials and probes. These were then averaged and the proportions of “long” responses for each stimulus interval were computed for each window of 100 trials moving forward in ∼60-second (∼25-trial) increments. The robustness of this analysis was further assessed across different window lengths: 50, 75, 125 and 150 trials (Supplemental materials). A GLMM (probit link) was then fitted to the proportion of long responses, including tonic DA concentrations and demeaned stimulus intervals and their interaction as fixed-effects parameters and by-stimulus slopes and intercepts for patient probes as random effects. As described above, this method yielded psychometric functions with separate beta coefficients reflecting temporal accuracy and precision (*41*). The windows were additionally partitioned according to the average DA concentrations into low, medium, and high DA terciles, thereby enhancing our ability to visualize the results. Finally, all analyses were repeated with SE concentrations to assess the neurochemical specificity of our findings. The Bayesian prevalence estimate was not computed for this analysis due to the small number of trial-windows.

### Supplementary Text

#### Time perception in PD

We assessed two contrasting hypotheses for the role of tonic striatal DA in temporal accuracy. The first, grounded in prior pharmacological studies (*20*, *77*, *78*), predicted a positive relationship, and the second, given the absence of clear evidence in dopamine-depleted PD (*54*), predicted no association. We observed a trend for a leftward shift of a psychometric function denoting an overestimation trend (Fig. 2A) in both PD patients, *PSE_PD_*=-.03 s, *SE*=.03, [95% CI:-.08, .02], and controls, *PSE_CTRL_*=-.04 s, *SE*=.03, 95% CI [-.08, .01], which, in line with the second hypothesis, was similar across the groups, *β*=.10, *SE*=.33, 95% CI [-.58, .79]), *z*=.31, *p*=.76, with Bayesian evidence for the null hypothesis, *BF_10_* = .09.

We additionally hypothesised that lower tonic DA levels will correspond with poorer precision (*49*, *50*). As expected, temporal precision was poorer in PD patients, *JND_PD_*=.14, *SE*=.02, 95% CI [.08, .19], as indicated by higher values (i.e., flatter slopes; Fig. 2A), than in controls, *JND_CTRL_*=.09, *SE*=.01, 95% CI [.08, .10], *β*=2.44, *SE*=1.03, 95% CI [.33, 4.56]), *z*=2.37, *p*=.02, *BF_10_* = 1.03. These results underscore diminished temporal precision in PD patients, corroborating previous findings amid some contrasting evidence (*49*, *50*).

#### Caudal dopamine and serotonin transients and time perception

We next tested the prediction that the elevated striatal DA transients are linked to temporal underestimation, as suggested by animal research (*25*). Toward this end, we investigated how patients’ neurochemical signals tracked variations in their temporal responses. Lower caudal DA after stimulus onset [625 to 670 ms] was associated with increased tendency to judge stimulus intervals as long, cluster *p*_TFCE_ <0.05 (Fig. 1C). This effect was dopamine-specific with no clusters found in SE time series (Fig. 1D). Importantly, the cluster analyses did not yield any statistically significant differences in DA or SE concentrations as a function of actual stimulus interval (modelled as both a binary variable [Fig. 1A,B] and with six stimulus levels). Finally, no clusters were identified in signal timeseries aligned to stimulus offset. Cumulatively, these results suggest that short responses are associated with larger bursts of DA activity during stimulus processing.

A complementary GLMM analysis corroborated that lower phasic DA at 625 – 670ms from stimulus onset was associated with increased long responses, *β*=-.11 (95% CI [-.22, -.001]), *SE*=.06, *t*=-1.97, *p*=.048, whereas SE was not a significant predictor of temporal estimates in this time window, *β*=-.06 (95% CI [-.16, .05]), *SE*=.05, *t*=-1.07, *p*=.28. This effect is further reflected in our analysis of a psychophysical measure of temporal accuracy (PSE): lower DA was associated with a greater overestimation bias, *β*=-.15 (95% CI [-.27, -.04]), *SE*=.06, *z*=-2.58, *p*=.01 whereas this effect was not significant for serotonin, *β*=-.01 (95% CI [-.13, .10]), *SE*=.06, *z*=-.22, *p*=.82. By contrast, temporal precision (JND) did not differ across phasic DA terciles, *β*=.34 (95% CI [-.43, 1.13]), *SE*=.40, *z*=.86, *p*=.39, or SE terciles, *β*=.61 (95% CI [-.18, 1.42]), *SE*=.41, *z*=1.51, *p*=.13 (Fig. 1E-J). These results complement the previous analysis and demonstrate that intra-individual variability in timing accuracy is related to transient caudal DA fluctuations, with evidence for neurochemical specificity.

To gain a deeper insight into these effects, we used a Bayesian approach to estimate population prevalence. This approach offers several benefits compared to traditional population mean hypothesis testing, including the capability to infer population-level estimates and their precision in studies with small participant numbers and to derive an estimate irrespective of the specific window where classification may peak in different participants (*42*). The AUC for classification of short and long responses from DA concentrations within individual participants varied from .50 to .59 (59% classification accuracy), reflecting heterogeneity across patients (Fig. S2 top-left). For example, whereas patient 20’s AUC ranged from .50 to .53, reflecting poor classification, patient 19 displayed the highest AUC values (.55<AUC<.60) around anchor stimulus interval offsets (500 and 1100 ms). The Bayesian estimate of population prevalence based on classification performance peaked at 47.38% at 620ms, which corresponds with our earlier results (Fig. 1). The Bayesian maximum a posteriori (MAP) (*79*) estimate of the effect prevalence was 82.50% (96% highest posterior density interval [HPDI]: [42.50, 98.80]) with uncorrected within-subject threshold α=.05 (Fig. S2 top-right). This suggests that there is at least a 42.50% chance of observing statistically significant classification of temporal judgments from DA signals within 1100ms from stimulus onset in a new sample if our methods are replicated (the prevalence estimate reduces to 32.70% with a more stringent threshold α=.01; Fig. S2 bottom-right).

**Figure S2.**
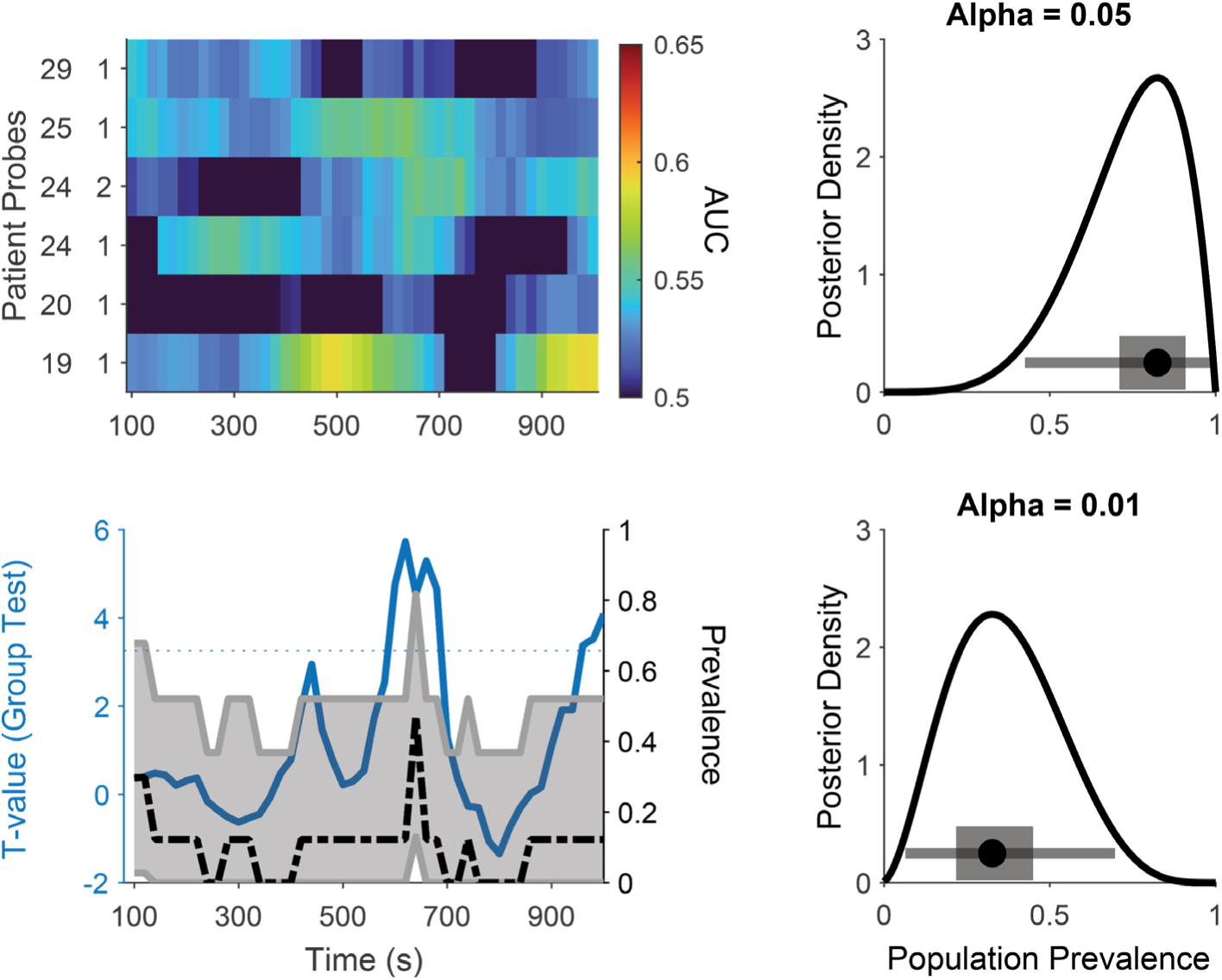
Bayesian population prevalence for classification of short and long responses from dopamine timeseries. (**Top-left**): Heatmap of AUC values for each patient probe (rows) and time window (200ms width, centred on x-axis labels). The blue solid line in **bottom-left** panel tracks the group-level t-test values, and the dotted blue line denote the threshold at α=0.01. The posterior distribution of population prevalence is shown for an effect in the analysed time series (cumulative density function in **right panels**) and at each timepoint (dashed black line in **bottom-left panel**). (**Right panels**) The grey horizontal line represents 96% highest posterior density intervals (HPDI), and the short rectangle shows the 50% HPDI.

#### Tonic caudal dopamine and serotonin concentrations and time perception

Our next set of analyses evaluated the association between within-subject variations in interval timing and *tonic* caudal DA levels. We contrasted the competing predictions: one suggesting that tonic DA is unrelated to temporal accuracy, given inconclusive evidence for atypical temporal bias in PD (*54*), and the other proposing a positive correlation, supported by pharmacological evidence (*22*, *80–82*). Consistent with the first hypothesis and our behavioural observations, tonic DA was not associated with temporal bias, *β*=-.004 (95% CI [-.06, .05]), *SE*=.03, *z*=-.14, *p*=.89, *BF_10_* = .06, underscoring that temporal accuracy is not related to steady-state (tonic) caudal DA levels (Fig. 2D,E). This finding stands in stark contrast to the compelling link observed for phasic DA levels. Furthermore, we also observed no association between tonic SE and temporal bias, *β*=-.04 (95% CI [-.12, .03]), *SE*=.04, *z*=-1.17, *p*=.24, *BF_10_* = .11 (Fig. 2G,H).

We additionally predicted that lower tonic DA levels, prevalent in PD, will correspond with poorer precision (*49*, *50*). Our behavioural data analysis supported this hypothesis, revealing poorer precision for patients than controls. Crucially, in our patient sample, temporal precision showed a positive association with their tonic DA concentrations (as shown by steeper slopes for high DA terciles in Fig. 2D), *β*=.50 (95% CI [.13, .87]), *SE*=.19, *z*=2.61, *p*=.01, *BF_10_* = 1.70, although the Bayesian evidence was not conclusive. We did not find a similar association with serotonin levels (Fig 2G), *β*=.26 (95% CI [-.28, .80]), *SE*=.27, *z*=.94, *p*=.35, *BF_10_* = .09. These findings support the hypothesis that fluctuations in tonic DA, but not SE, are linked to temporal precision. As observed earlier for temporal accuracy, the results for temporal precision further emphasize the contrast in the impact of tonic vs. phasic dopamine concentrations on interval perception, highlighting potentially differential effects of distinct temporal dynamics of DA activity.

